# Clustering and longitudinal change in SARS-CoV-2 seroprevalence in school-children: prospective cohort study of 55 schools in Switzerland

**DOI:** 10.1101/2020.12.19.20248513

**Authors:** Agne Ulyte, Thomas Radtke, Irene A. Abela, Sarah R. Haile, Christoph Berger, Michael Huber, Merle Schanz, Magdalena Schwarzmueller, Alexandra Trkola, Jan Fehr, Milo A. Puhan, Susi Kriemler

**Affiliations:** Epidemiology, Biostatistics and Prevention Institute (EBPI), University of Zurich, Hirschengraben 84, 8001 Zürich, Switzerland; Institute of Medical Virology, University of Zurich, Winterthurerstrasse 190, 8057 Zürich, Switzerland; University Children Hospital Zurich, Steinwiesstrasse 75, 8032 Zürich, Switzerland; Epidemiology, Biostatistics and Prevention Institute (EBPI), University of Zurich, Hirschengraben 84, 8001 Zürich; Division of Infectious Diseases & Hospital Epidemiology, University Hospital Zurich, Rämistrasse 100, 8091 Zurich, Switzerland

**Keywords:** SARS-CoV-2, COVID-19, children, adolescents, school

## Abstract

**Background and aims:** The facilitating role of schools in SARS-CoV-2 infection spread is still debated and the potential of school closures to mitigate transmission unclear. In autumn 2020, Switzerland experienced one of the highest second waves of the SARS-CoV-2 pandemic in Europe while keeping schools open, thus offering a high-exposure environment to study SARS-CoV-2 infections in schools. The aim of this study was to examine longitudinal change in SARS-CoV-2 seroprevalence in children and the evolution of clustering within classes and schools from June to November, 2020, in a prospective cohort study of school children in the canton of Zurich, Switzerland.

**Methods:** Children from randomly selected schools and classes, stratified by district, were invited to participate in serological testing of SARS-CoV-2 in June-July and October-November 2020. Parents of children filled questionnaires on sociodemographic and health-related questions. 55 schools and 275 classes within them were enrolled, with 2603 children participating in the first, and 2552 in the second testing (age range 6-16 years). We evaluated longitudinal changes of seroprevalence in districts and investigated clustering of seropositive cases within classes and schools.

**Results:** Overall SARS-CoV-2 seroprevalence was 2.4% (95% CrI 1.4%-3.6%) in summer and 4.5% (95% CrI 3.2%-6.0%) in not previously seropositive children in late autumn, leading to estimated 7.8% (95% CrI 6.2%-9.5%) of ever seropositive children, without significant differences among lower, middle and upper school levels. Among the 2223 children with serology tested twice, 28 (40%) of previously seropositive were negative, and 109 (5%) previously negative became seropositive. Seroprevalence was not different between school levels or sexes, but varied across districts (1.7% to 15.0%). Between June-July and October-November 2020, the ratio of diagnosed to newly seropositive children was 1 to 8. At least one newly seropositive child was detected in 47 of 55 schools and 90 of 275 classes. Among 130 classes with high participation rate, 0, 1-2 or ≥3 seropositive children were present in 73 (56%), 50 (38%) and 7 (5%) classes, respectively. Class level explained slightly more variation of individual serological results (standard deviation of random effects (SD) 0.97) than school level (SD 0.61) in the multilevel logistic regression models. Symptoms were reported for 22% of seronegative and 29% of newly seropositive children since summer.

**Conclusions:** Under a regimen of open schools with some preventive measures in place since August, clustering of seropositive cases occurred in very few classes and not across entire schools despite a clear increase in seropositive children during a period of high transmission of SARS-CoV-2.

**Trial registration:** ClinicalTrials.gov NCT04448717. https://clinicaltrials.gov/ct2/show/NCT04448717

## Introduction

Severe acute respiratory syndrome coronavirus 2 (SARS-CoV-2) infection and the role of schools is still a controversy: children, particularly adolescents, can be infected as often as adults [1,2] but rarely develop clinically manifest coronavirus disease 2019 (COVID-19) or severe health outcomes [3–6]. However, the prevalence of asymptomatic and oligosymptomatic infections and potential spread in schools is still debated. School closures have been implemented in many countries in the first half of 2020 in order to curb the pandemic, leading to disrupted education for 1.5 billion learners in up to 172 countries in April 2020 [7]. On December 17, more than 0.3 billion learners were still affected by school closures [7].

The negative effects of school closures include an increase in social and economic inequality, and adverse long-term educational, social and health outcomes for children [8– 10]. There is less uncertainty about the mentioned negative effects of school closures than about the risks that school setting presents for transmission of SARS-CoV-2. However, evidence has accumulated that children in schools and households are not the main drivers of the infection spread [11–13].

Although a few outbreaks in educational settings have been reported worldwide during early 2020 (e.g., in Israel [14], the US [15]), full or partial opening of schools in many countries in Autumn 2020 has not been followed by more frequent outbreaks [16–19].

It is thus unclear how frequent silent or noticeable outbreaks in classes and entire schools are. Most studies of SARS-CoV-2 in schools have focused on identified SARS-CoV-2 cases and subsequent contact-tracing within schools. Thus, asymptomatic or oligosymptomatic children infected with SARS-CoV-2 are still likely missed and it is not clear how frequent and significant they are for infection spread. There is a need for longitudinal, population and school-based studies with random sampling on class and school level.

In this article, we present the results of the longitudinal cohort study *Ciao Corona* in the canton of Zurich, Switzerland with measurement of SARS-CoV-2 antibodies and symptoms in a cohort of more than 2500 children from 55 schools in June-July and October-November 2020 (hereafter referred to as T1 and T2). As for most of Europe, schools were open in this most populous canton of Switzerland since late August 2020, with preventive measures in place. Switzerland experienced one of the highest second waves of the SARS-CoV-2 pandemic in Europe in autumn 2020, thus presenting a high-exposure environment to study SARS-CoV-2 infections in schools. *Ciao Corona* is one of the few prospective, population and school-based studies of SARS-CoV-2 seroprevalence, and takes place in a country with one of the highest SARS-CoV-2 incidences worldwide, offering unique insights into the change in clustering of seropositive cases within classes and schools, and the association with self-reported symptoms. The aims of the study were to estimate the longitudinal change of seroprevalence, clustering within schools and classes, and to assess the association with reported symptoms.

## Methods

This longitudinal cohort study is registered on ClinicalTrials.gov (identifier number NCT04448717) and the protocol is reported elsewhere [20]. The study is part of the nationally coordinated research network in Switzerland, *Corona Immunitas* [21]. The canton of Zurich, in which the study was based, comprises 1.5 million linguistically and ethnically diverse residents, approximately 18% of Swiss population, residing both in urban and rural settings. In 2020, physical attendance of schools was interrupted only between March 16 and May 10. Implemented preventive measures (e.g., masks for school personnel and children in secondary schools, reduction of some large group activities) have fluctuated since then. However, the schools have been in continuous operation since the start of the school year on August 17 to the end of 2020. Once an index case was identified in a school, children and school personnel were quarantined based on contact-tracing of close contacts. Full classes were quarantined only when two or more infected students were identified within a class. All schools were obliged to develop a plan and implement specific preventive measures by August (e.g., masks for teachers and children >12-years-old, distancing rules in class- and teachers’ rooms, tapering of school breaks, no mixing of classes, ban of group gatherings such as excursions and camps beyond class units, no parents on school grounds) to mitigate transmission but they varied from school to school. Common to all schools was the requirement to keep children at home if they are sick beyond very mild symptoms such as runny nose or mild cough, masking for adults in the school from 19 October and additionally for children of secondary schools (above 12-years-old) from 2 November.

### Population

Primary schools were selected randomly from the list of all schools in the canton of Zurich, stratified by region, and the geographically closest secondary school (often, in the same school building) matched. From 156 invited, 55 schools agreed to participate. Classes within participating schools were selected randomly, stratified by school level: grades 1-2 in lower level (attended by 6 to 9-year-old-children), grades 4-5 in middle level (attended by 9 to 13-year-old children) and grades 7-8 in upper school level (attended by 12 to 16-year-old-children). Invited grades were selected to ensure that the same cohort of children will remain in the classes until April 2021 (children in grades 3, 6 and 9 often change the class and school in the next year). We aimed to enroll at least three classes and at least 40 children in each school level at the invited schools. Major exclusion criterion was suspected or confirmed SARS-CoV-2 infection in the given child during the testing (precluding the attendance of the testing at school).

First round of testing (T1) was completed with 2585 participants (serology results available for 2484) in June-July 2020, and first results are reported elsewhere [1]. 18 eligible children, who could not participate in June-July, were additionally tested in August (serology results available for 12), and results merged to T1 testing round, resulting in a total of 2603 children at T1. Flowchart of study participants is shown in Figure 1.

**Figure 1.**
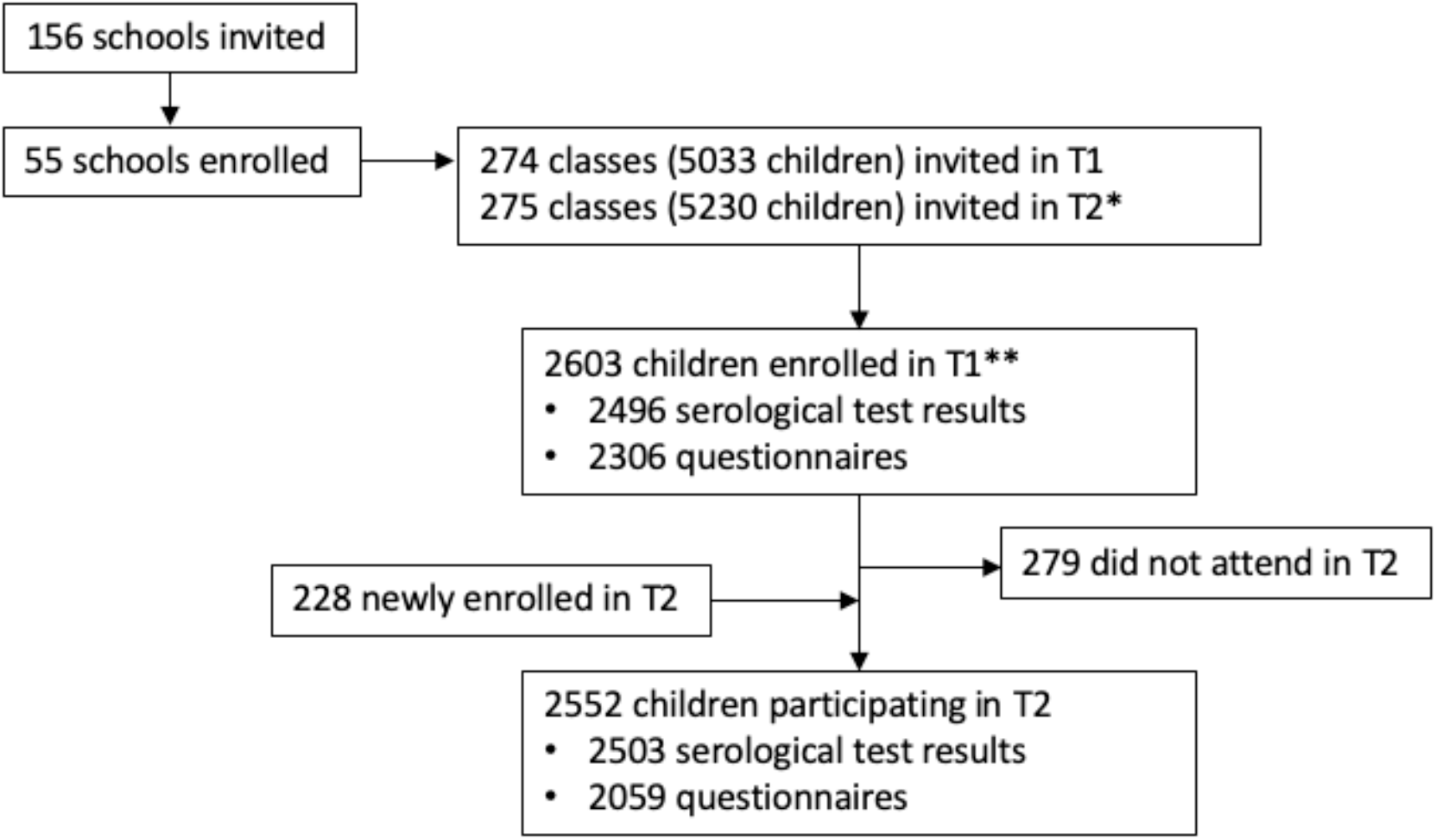
Flowchart of the cohort study participants T1 – testing in June-July 2020. T2 – testing in October-November 2020. *Some classes were split or rearranged into multiple classes after the summer break. **18 of these children were enrolled in August/September (12 serological results, 18 questionnaires).

The same cohort of classes was invited to the second testing round (T2) in October-November. If the previously invited class had been restructured after the summer break, all children in the newly formed classes attended by previously enrolled participants were eligible to participate.

### Serological testing

Collection of venous blood samples was performed in participating schools in October 26 – November 19, 2020.

Venous blood samples were analyzed with the ABCORA 2.0 binding assay of the Institute of Medical Virology (IMV) of the University of Zurich, which is based on Luminex technology. The test was described in detail in previously [1]. The test analyzes immunoglobulins G (IgG), M (IgM) and A (IgA) against four SARS-CoV-2 targets (receptor binding domain (RBD), spike proteins S1 and S2, and the nucleocapsid protein (N), yielding 12 different measurements. Cut-off values were established against pre-pandemic plasma allowing a high sensitivity (94.3%) and specificity (99.0%) (see [22] for test description; test parameters have been updated since based on expanded validation cohort). Samples were defined as seropositive for SARS-CoV-2 if at least two of the 12 parameters were above the cut-off.

Three serological outcomes were analyzed, combining the results at T1 and T2. First, we estimated seroprevalence in June-July (T1) among all children tested. Second, we estimated seroprevalence in October-November among previously non-positive or non-tested children since June-July (T2). Even though some of the seropositive children at T1 tested negative at T2, we excluded them from T2 results because of a potentially persisting cellular immunity [23] and generally rare reports of reinfection in children, and thus a low likelihood of a repeated infection since summer. Third, we estimated the proportion of children who ever had a SARS-CoV-2 infection based on serological test results by October-November by analyzing the children tested at T2 (or seropositive at T1 and not retested at T1) and counting as positive those who were tested positive at T1 (regardless of subsequent T2 results) or T2. Seroprevalence outcomes are summarized in Table 1.

**Table 1.** Definitions of seroprevalence outcomes

| Abbreviation | Definition | Numerator | Denominator |
| --- | --- | --- | --- |
| T1 seroprevalence (seropositive in summer testing) | Seroprevalence in children in June-July 2020 | Seropositive children in June-July | All tested children in June-July* |
| T2 seroprevalence (newly seropositive in autumn testing) | Seroprevalence in children, susceptible at June-July, in October-November 2020 | Seropositive children in October-November, excluding those who also tested seropositive in June-July | All tested children in October-November, excluding those who also tested seropositive in June-July |
| T1+T2 seroprevalence (ever seropositive in summer and/or autumn) | Proportion of children who had been infected with SARS-CoV-2 by October-November 2020, as reflected by ever being seropositive | Children tested seropositive at least once in June-July or October-November (ever tested seropositive) | All tested children in October-November and seropositive children tested in June-July who did not participate in the testing in October-November |

\* also including 18 children tested (serological results available for 12) in late August-early September

### Statistical analysis

Statistical analysis included descriptive statistics and Bayesian hierarchical modelling to estimate seroprevalence, accounting for the sensitivity and specificity of the SARS-CoV-2 antibody test, the hierarchical structure of cohort (individual and school levels), and post-stratification weights, which adjusted for the population-level grade level at school and geographic district [2]. The factor of confirmed to total infections (dark figure) was calculated as the ratio of real-time polymerase chain reaction (rtPCR) confirmed cumulative incidence of SARS-CoV-2 infections from July 30 (after the first testing was completed) to November 8 2020 (during autumn testing period) and the total cumulative incidence until November 8, based on official statistics [24], and the estimated autumn (T2) and overall (T1+T2) seroprevalence.

Clusters within classes were defined as three or more cases of newly seropositive children within a class in autumn (T2) testing. We estimated the proportion of classes with a cluster of seropositive children by dividing their number by the total number of enrolled classes where at least 5 children and at least 50% of the children were tested at the relevant time point. This way, classes with low participation rate and very small classes (less than 5 children) were excluded from the analysis of clusters, as likely to be misclassified in the cluster analysis. However, if a cluster was identified in a class with less than 50% participation rate, the class was additionally included both in the numerator and the denominator for the proportion of classes with clusters. Clustering of seropositive children within classes was further examined by comparing our study results with a simulation of a hypothetical study with the population (classes and numbers of children tested at T2) identical to this study. In the simulation, independent chance of seropositive results (i.e., no association of seropositive cases within a class) was assumed, equal to the observed proportion of T2 seropositive among all T2 tested children. By comparing the number of classes with clusters actually observed in our study and that in the simulation, we could estimate if such number of clusters would be likely to be observed by chance.

Semi-structured interviews with the principals of schools with classes with observed clusters of T2 seropositive children were performed after T2 testing to further investigate the detected clusters. Interview questions covered numbers of diagnosed and quarantined teachers and children in the affected classes, potential temporal sequence of infections and other related circumstances.

To determine whether schools or specific classes explained more of the variance in seropositivity, individual level serology results were modeled in a multilevel logistic regression, with sex and school level (as a proxy for age) as fixed effects. Three models were compared: with random effects for school level, with random effects for class level, and with both random effects.

Data analysis was performed with R version 4.0.3 [25]. Bayesian hierarchical modelling was performed using the R package rstan [26].

## Results

In total, 2831 children from 275 classes within 55 schools in the canton of Zurich were enrolled in the study by October-November 2020. From these, 2603 participated in T1 summer testing, and 2552 participated in T2 autumn testing. The flowchart of enrolled participants with serological test results and questionnaire information available is shown in Figure 1.

At T2 testing, serological results were available for 731 children from lower school level (median age 8, age range 6-10 years), 863 children from middle school level (median age 11, age range 8-13 years), and 909 children from upper school level (median age 14, age range 11-16 years). 1287 children were female, 1211 male and 5 reported other gender. Median participation rate at T2 within a class was 47% (interquartile range 30%-62%).

Serological test results were positive for 74 children in T1 testing. For these children, in T2 testing serology result was positive for 42 (60%, median age 10 years, age range 7-14 years, symptoms reported in 31/41 (76%) in January-July) and negative for 28 (40%, median age 10 years, age range 7-14 years, symptoms reported in 17/24 (71%) in January-July), while 4 did not attend T2 testing. Serological results were positive for 173 children in T2, including 109 children who tested negative in T1, and 22 newly enrolled children. The complete distribution of positive results in T1 and T2 is shown in the Appendix1.

Estimated SARS-CoV-2 seroprevalence in T1 in children was 2.4% (95% CrI 1.4%-3.6%). Seroprevalence in autumn (newly seropositive in T2) in children was 4.5% (95% CrI 3.2%-6.0%) (Figure 2). The proportion of children having had SARS-CoV-2 infection (ever seropositive, T1+T2) by autumn was 7.8% (95% CrI 6.2%-9.5%). The seroprevalence at T1 and T2 does not add up to the proportion of ever seropositive children (T1+2) as the populations included in the numerator and denominator of these three outcomes are not exactly the same (see Table 1 and explanation in Appendix2). The range of newly seropositive children in the districts of the canton of Zurich was 1.7%– 15.0%, and the range of proportion of ever seropositive children 3.5%-21.2% (Figure 2). T2 seroprevalence in lower, middle and upper school levels was 4.4% (95% CrI 2.7-6.7%), 5.0% (95% CrI 3.0-7.4%) and 3.9% (95% CrI 2.1-6.2%), respectively, and T1+T2 seroprevalence in lower, middle, and upper school level was 8.5% (95% CrI 6.1-11.4%), 8.0% (95% CrI 5.7-10.7%) and 6.4% (95% CrI 4.3-8.9%), respectively. The difference in estimated seroprevalence was not different between males and females (T2: 4.8% vs 4.2%; T1+2: 8.3% vs 7.2%).

**Figure 2.**
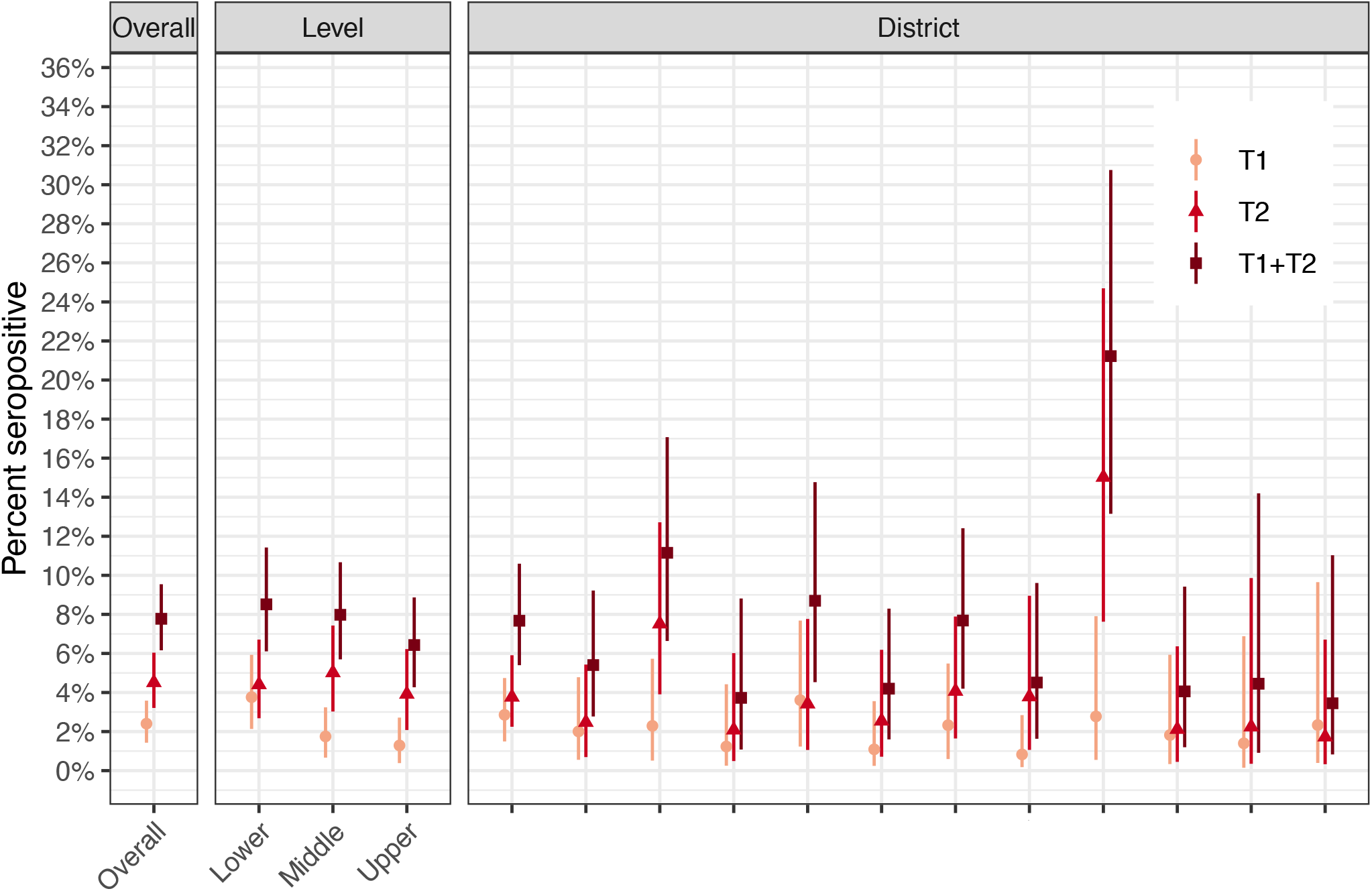
Seroprevalence estimates in school levels, sexes and districts of the canton of Districts are ranked in the order of decreasing population size.

Compared to the cumulative incidence of SARS-CoV-2 infections between July and November in the canton of Zurich in children between 4 and 15-years-old, the ratio of diagnosed to seropositive children was 1 to 8. The ratio of total cumulative incidence of SARS-CoV-2 infections in children since January to November 2020 to ever seropositive children was 1 to 13.

The number of newly T2 (ever T1+T2) seropositive children within a school-level ranged from 0 to 12 (0 to 14), and within a class from 0 to 10 (0 to 11). At least one newly seropositive child was detected in 47 out of 55 schools and in 90 out of 275 classes (56 out of 129 classes with ≥5 children and ≥50% of children tested; 57 out of 130 classes with high participation rate or at least three newly seropositive children).

At least one ever seropositive child was detected in 52 out of 55 schools and in 125 out of 275 classes (75 out of 129 classes with ≥5 children and ≥50% of children tested). Distribution of newly and ever seropositive children within tested classes with more than 5 children and more than 50% of the class tested is shown in Figure 3.

**Figure 3.**
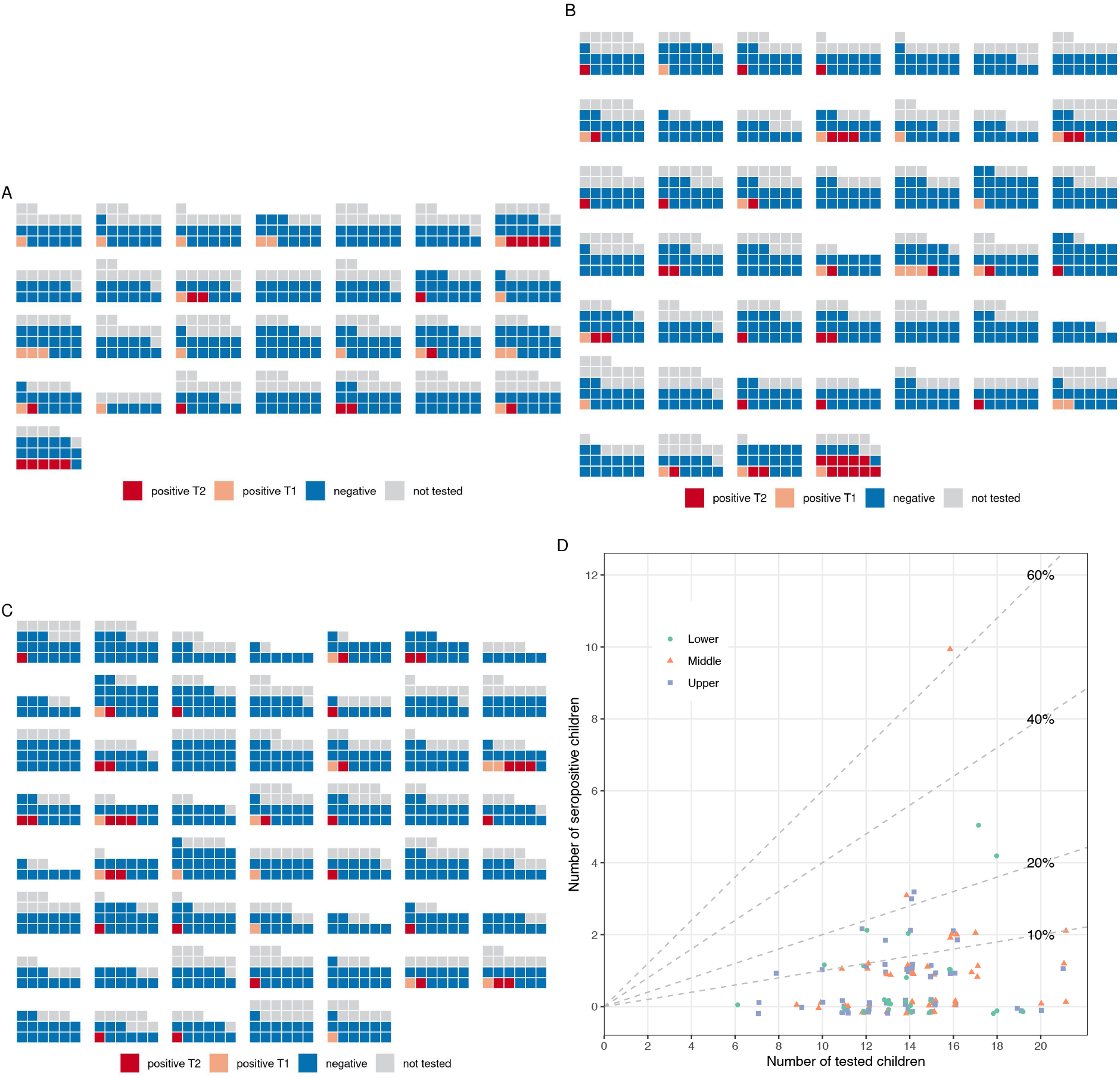
Distribution of children seropositive at T1 and newly at T2 in classes with ≥5 and ≥50% of children tested Figure depicts children tested at T2 (autumn 2020): seronegative (blue), seropositive in T1 (orange), newly seropositive in T2 (red), and not tested or without serological results available in T2 (grey). A – lower school level classes, B – middle school level classes, C – upper school level classes. D presents the summary of the newly seropositive children at T2 numbers in the classes depicted in A-C.

Seven classes in five schools had three or more newly seropositive children: three classes in lower, two in middle, and two in upper school levels. Detailed information about the clusters is provided in Table 2. Assuming a uniform 5.4% seropositivity rate across all tested children, and numbers of children tested within classes as observed in this study, a simulation study showed that 7 or more clusters would be expected by chance in 14% of repetitions, with median expected number of classes with such clusters 4 (95% CrI 1 – 9). Thus, even if infections within classes were not associated, in a population with the classes structure and total number of seropositive children as in this study, we would expect to see 4 clusters of three of more seropositive children in a class.

**Table 2.** Details about the classes with three or more newly seropositive children between July and November 2020

| School |  | N of children in the class |  |  | PCR positivity |  | Quarantine/Isolation |  | Information | Probable index case |  |  |
| --- | --- | --- | --- | --- | --- | --- | --- | --- | --- | --- | --- | --- |
| ID | Level | Total | Tested | Newly seropositive | Teacher | Children | Teacher | Children |  | Teacher | Child | Household |
| 1 | middle | 17 | 14 | 3 | 0 | 0 | 0 | 8 | Individual unrelated cases in quarantine due to positive household members. |  |  | x |
| 2 | lower | 19 | 9 | 3 | 0 | 0 | 0 | 0 | Cases not previously known. | ? | ? | ? |
| 3 | lower | 25 | 16 | 4 | 1 | 2 | 1 | Whole class | Teacher tested positive; next day 2 children tested positive. | x |  |  |
| 4 | medium | 23 | 16 | 10 | 1 | 3 | 1 | Whole class | Child tested positive; next day 2 other children and teacher positive; many household members of the class infected as well; index case unclear. | ? | ? | ? |
|  | lower | 22 | 17 | 5 | 0 | 0 | 0 | 2 | Two unrelated children in quarantine due to diagnosed sports camp coach and affected household member. |  |  | x |
| 5 | upper | 17 | 13 | 3 | 0 | 1 | 0 | Whole class | Class quarantined after a mother and subsequently the child (class student) tested positive, while the class was in a camp. |  |  | x |
|  | upper | 16 | 12 | 3 | 0 | 0 | 0 | 3 | Individual unrelated cases (one due to infected brother in another class, other two unknown). |  | ? | ? |

In the multilevel logistic regression models of individual serology results of T2 serology, school level (as a proxy for age) and sex were not significant predictors. Estimated standard deviation (SD) of random effects for the school decreased once the random effect for class was added (from 0.606 (95% CI 0.400-0.864) to 0.467 (95% CI 0-0.786)). Estimated SD of random effects for class remained relatively stable and was bigger than the effect of school once the random effect of school was added (from 0.969 (95% CI 0.692-1.290) to 0.767 (95% CI 0.371-1.170)).

Symptoms between the summer break and November 2020 were reported in 21.8% seronegative and in 28.7% newly seropositive children (T2). The distribution of individual symptoms is depicted in Figure 4. Although reported rarely in general, only loss of smell or taste was more frequent in seropositive than in seronegative children, (3/101 (3.0%) vs 4/1923 (0.2%)). The most frequently reported symptoms in seropositive children were headache (13.9%), runny or congested nose (11.9%), sore throat (11.9%), and fatigue (8.9%).

**Figure 4.**
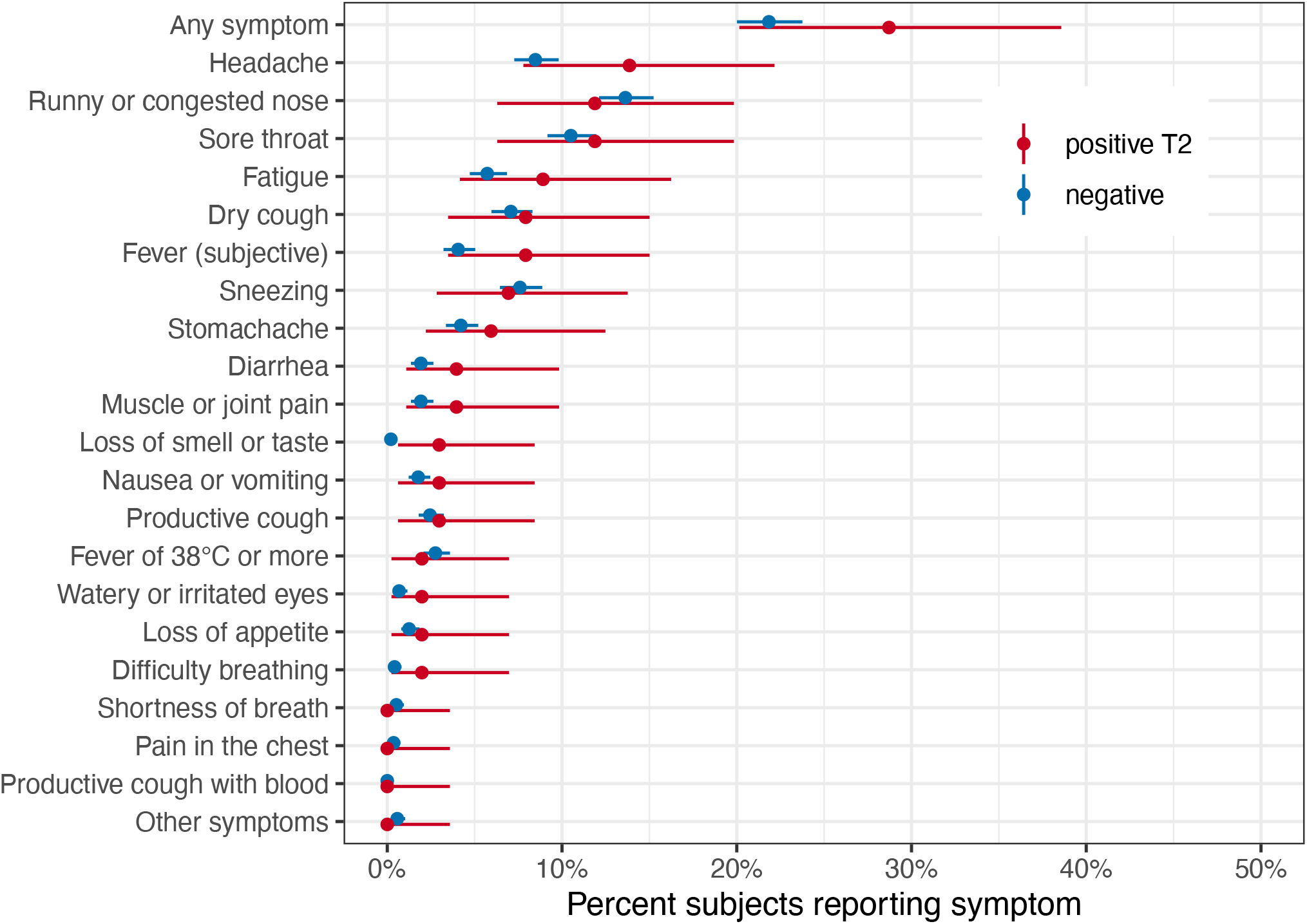
Symptoms reported between July and November 2020 in seronegative and newly seropositive (T2) children

## Discussion

In this cohort study of 55 schools and more than 2500 children, we observed only minimal clustering of seropositive cases within classes and schools between July and November 2020 despite a clear increase in seroprevalence among children during a time of very high transmission. Clusters could be partially explained by independent occurrence of cases from individual infections among classmates given the high community transmission of SARS-CoV-2. SARS-CoV-2 seroprevalence increased since June-July from 2.4% to 7.8% in October-November. SARS-CoV-2 antibodies were not detected after four months for 40% previously seropositive children. There was no difference in seroprevalence between school levels (age groups), although a trend of lower seroprevalence was observed in older children.

In autumn 2020, Switzerland had one of the highest reported incidences of SARS-CoV-2 infections in Europe, peaking at approximately 950 daily cases per million inhabitants in early November [27]. A similarly high incidence was observed in the canton of Zurich, where the study was conducted, with approximately 590 daily cases per million inhabitants and rtPCR test positivity of approximately 16% recorded for the first half of November [24]. However, schools have been open since the school year started on August 17, 2020. Some but not extensive preventive measures were implemented, such as masks for school personnel and restriction of large group activities. As the number of cases in the community increased, masking for children in secondary schools (over 12-years-old) was implemented from November. In light of this context, the increase in seroprevalence since summer is expected. However, it was not accompanied by high incidence of clustered seropositive cases within schools and classes. This finding documents the potential for schools to remain open and operate safely, without substantial risk for frequent outbreaks, even in a high community transmission setting.

Clusters of 3 or more newly seropositive children were observed only in the minority of classes (six out of 129 classes with high participation rate, and in one class with 47% participation rate). In contrast to some other studies, we did not observe higher seroprevalence or clustering of cases in children of older age in the secondary schools. Potentially, behavioral factors and preventive measures in schools including masking of children in higher school grades helped to mitigate the potential spread of infection. The ratio of diagnosed to seropositive cases among children, although still alarming, had decreased substantially since summer, from 1 to 89 [1] to 1 in 8 cases, meaning that diagnosis of the infected children had dramatically improved.

Observed clustering of cases within a class does not necessarily signal an outbreak (internal infection spread) has occurred in the school. In the seven classes with observed potential clusters, seropositive children were likely not part of the same infection transmission chain in at least two of the classes. In six classes, at least some of the seropositive children were previously diagnosed or quarantined. The results of the simulation showed that even if seropositive status was assigned to children of the study population completely randomly, clusters of seropositive children would be observed in seven or more of the tested classes with 14% probability. Thus, even if the seropositive children were not associated within classes (i.e., infections completely independent of each other), it would be not unreasonable to expect to see as many clusters as observed in this study just by chance.

When clustering did exist, it seemed to be related to the class rather than to the school, as suggested by the multilevel models. This could mean that, as could be expected, infection is more likely to spread within a class rather than school (if at all). Potentially, the random effect of the school would become even smaller once the incidence in the community (district) is controlled for. This is another reason why focused class-based quarantine measures may be more efficient than penalizing whole schools or even the entire school system for localized clustering.

This study offers unique insights into the transmission and prevalence of SARS-CoV-2 infection in schools on a randomly selected, representative, longitudinal, population level cohort. Most of the other studies of SARS-CoV-2 in schools have focused on contact tracing of index cases [16,28], thus potentially missing unidentified cases. Other studies have relied on the prevalence of diagnosed SARS-CoV-2 cases to estimate the frequency of outbreaks and risk for children infected while schools are operating in person [16–18]. Finally, a few ecological studies tried to estimate the overall effect of closing and opening schools on the development of the pandemic (in terms of diagnosed and reported cases and deaths) [29– 31], with major limitations of uncontrolled confounding, high level of aggregation (e.g., pooling school and university closures as one intervention, or analyzing aggregated outcomes on country-level) and potentially measuring the outcome in a population not exposed to the intervention. Finally, a stochastic modelling study of infection spread in schools have shown that some, although minimal, clustering of infections (outbreaks) is likely to happen even if major prevention and screening strategies are implemented [32].

In contrast to the mentioned retrospective and modelling studies, our study offers a prospective population level view, corresponding to school structure thanks to sampling on the school and class levels. In addition, having measured the baseline seroprevalence in June-July 2020, we were able to study the incidence of newly seropositive cases and their clustering in classes and schools in autumn. The study had a very high retention rate, with 89% of enrolled children retested in autumn. Together with the newly enrolled children from the same classes joining in autumn, the study had a high overall participation rate, especially given that it included venous blood sampling in children. High participation rate within a large proportion of classes allowed to study clustering on class level, which has not been possible in other (rare) seroprevalence studies in children [33].

The study has a few limitations. First, seroprevalence does not necessarily reflect the exact levels of past infection. Although we were able to adjust for test accuracy parameters on the population levels, leading to accurate estimates of overall seroprevalence, some false negatives and positives can be expected on individual level. In comparison to the summer testing, the number of false positives is expected to be lower as the prevalence has increased [34]. Among the 131 newly positively tested children in autumn, 20 would be expected to be false positive and among the 2330 negatively tested, 11 false negative, based on the estimated seroprevalence and test accuracy parameters. This means that the true rate of clustering could be expected to be even lower. Second, measuring seroprevalence rather than acute diagnosis of SARS-CoV-2 allows only a retrospective analysis, and prevents full reconstruction of the temporal sequence of infected cases within classes. In addition, seroprevalence is a dynamic parameter, as some children lose the antibodies and thus might appear seronegative despite having had the virus. Based on the very low re-infection rates in the literature, serological status only tells a partial story about the immunity against SARS-CoV-2 infections. Other unspecific or T-cell mediated cellular responses may exist to confer long-term immunity [23]. However, the limitation would be even higher with diagnostic testing, within significantly smaller window of positivity and no retrospective information. We were able to reconstruct at least some of the temporal information by comparing the serology status of children in summer and autumn, and thus differentiating cases infected in the first (spring) and second (autumn) wave of the pandemic, and by interviewing school principals about development of infections in classes.

In conclusion, clustering of cases occurred in very few classes and not across entire schools despite a striking increase in individual seroprevalence in children during a period of high transmission of SARS-CoV-2. As SARS-CoV-2 pandemic mitigation and the debate of the role of schools in the transmission of infections remains relevant, this study brings evidence that clusters of SARS-CoV-2 infection are rare in schools, and the transmission is more likely to be limited to classes. These findings from a country with high community transmission give hope that schools can be kept open, given that preventive measures are implemented within the school and the surrounding community.

## Ethics approval

The study was approved by the Ethics Committee of the Canton of Zurich, Switzerland (2020-01336). All participants provided written informed consent before being enrolled in the study.

## Data sharing statement

Data is still being collected for the cohort study *Ciao Corona*. Deidentified participant data might be available on reasonable request by email to the corresponding author at later stages of the study.

## Data Availability

Data is still being collected for the cohort study Ciao Corona. Deidentified participant data might be available on reasonable request by email to the corresponding author at later stages of the study.

## Funding

This study is part of *Corona Immunitas* research network, coordinated by the Swiss School of Public Health (SSPH+), and funded by fundraising of SSPH+ that includes funds of the Swiss Federal Office of Public Health and private funders (ethical guidelines for funding stated by SSPH+ will be respected), by funds of the Cantons of Switzerland (Vaud, Zurich and Basel) and by institutional funds of the Universities. Additional funding, specific to this study is available from the University of Zurich Foundation. The funder/sponsor did not have any role in the design and conduct of the study; collection, management, analysis, and interpretation of the data; preparation, review, or approval of the manuscript; and decision to submit the manuscript for publication.

---

**Appendix 1.**
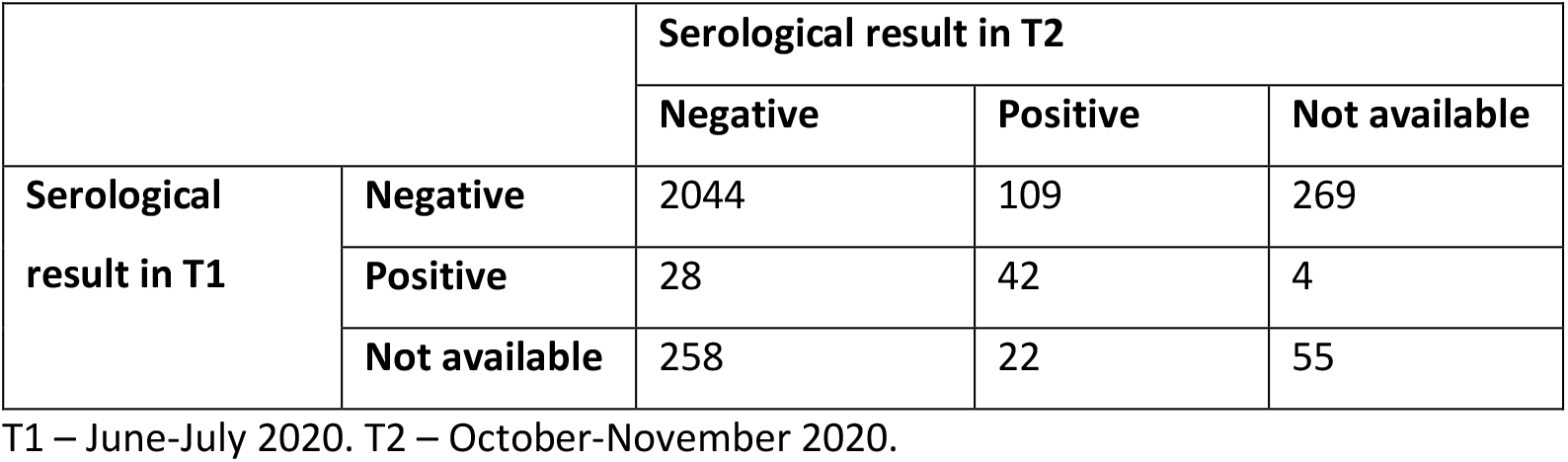
Comparison of individual serological results in study participants in summer (T1) and autumn (T2)

**Appendix 2.** Comparison of numerator and denominator in estimating seroprevalence at different time points of the study (T1, T2 and T1+T2)

| Serology test results |  |  |  | Analysis of seroprevalence |  |  |
| --- | --- | --- | --- | --- | --- | --- |
| T1 | T2 | T1+T2 | N | T1 | T2 | T1+T2 |
| - | - | - | 2044 | - | - | - |
| - | + | + | 109 | - | + | + |
| - | NA | NA | 269 | - |  |  |
| + | - | + | 28 | + | * | + |
| + | + | + | 42 | + | ** | + |
| + | NA | + | 4 | + |  | + |
| NA | - | - | 258 |  | - | - |
| NA | + | + | 22 |  | + | + |
| NA | NA | NA | 55 |  |  |  |
| Number of positive results (numerator) |  |  |  | 74 | 131 | 205 |
| Total number analyzed (denominator) |  |  |  | 2496 | 2433 | 2507 |
| Raw proportion of seropositive results |  |  |  | 3.0% | 5.0% | 8.2% |

N – number of participants in the corresponding row.
- negative result at the testing, + positive result at the testing, NA – results not available.
Cells in yellow are used for the analysis of seroprevalence at the corresponding time points.
\* previously seropositive results that are seronegative at T2 are not counted, as immunity might be still persisting, thus leading to a different further susceptibility and rates of infection than in those previously seronegative.
\*\* previously seropositive results that are still seropositive at T2 are not counted in T2, as they are more likely related to infections in spring (before T1) rather than in summer and autumn (between T1 and T2).
Raw proportion of seropositive results does not correspond to the seroprevalence estimated with Bayesian hierarchical models (see Methods and Results), which additionally corrects for test accuracy and population structure parameters. Estimated seroprevalence T1+T2 is higher than the sum of T1 and T2 seroprevalence – similarly as for the raw proportions of seropositive results, shown in this table – due to slightly different denominators used in the estimation at each time point.

## Notes

### Competing Interest Statement

The authors have declared no competing interest.

### Clinical Trial

NCT04448717

### Clinical Protocols

https://doi.org/10.1007/s00038-020-01495-z

## References

1 Ulyte A, Radtke T, Abela IA, et al. Variation in SARS-CoV-2 seroprevalence in school-children across districts, schools and classes. medRxiv 2020;:2020.09.18.20191254. DOI:10.1101/2020.09.18.20191254

2 Stringhini S, Wisniak A, Piumatti G, et al. Seroprevalence of anti-SARS-CoV-2 IgG antibodies in Geneva, Switzerland (SEROCoV-POP): a population-based study. Lancet 2020;0. DOI:10.1016/S0140-6736(20)31304-0

3 Levin AT, Hanage WP, Owusu-Boaitey N, et al. Assessing the Age Specificity of Infection Fatality Rates for COVID-19: Systematic Review, Meta-Analysis, and Public Policy Implications. medRxiv 2020;:2020.07.23.20160895. DOI:10.1101/2020.07.23.20160895

4 Shekerdemian LS, Mahmood NR, Wolfe KK, et al. Characteristics and outcomes of children with coronavirus disease 2019 (COVID-19) infection admitted to US and Canadian pediatric intensive care units. JAMA Pediatr 2020;174:868–73. DOI:10.1001/jamapediatrics.2020.1948

5 Hoang A, Chorath K, Moreira A, et al. COVID-19 in 7780 pediatric patients: A systematic review. EClinicalMedicine 2020;0:100433. DOI:10.1016/j.eclinm.2020.100433

6 Götzinger F, Santiago-García B, Noguera-Julián A, et al. COVID-19 in children and adolescents in Europe: a multinational, multicentre cohort study. Lancet Child Adolesc Heal 2020;4:653–61. DOI:10.1016/S2352-4642(20)30177-2

7 UNESCO. School closures caused by Coronavirus (Covid-19). 2020. https://en.unesco.org/covid19/educationresponse (accessed 19 Dec 2020).

8 Lancker W Van, Parolin Z. COVID-19, school closures, and child poverty: a social crisis in the making. Lancet Public Heal 2020;5:e243. DOI:10.1016/S2468-2667(20)30084-0

9 Norman RE, Byambaa M, De R, et al. The Long-Term Health Consequences of Child Physical Abuse, Emotional Abuse, and Neglect: A Systematic Review and Meta-Analysis. PLoS Med 2012;9:e1001349. DOI:10.1371/journal.pmed.1001349

10 Christakis DA, Van Cleve W, Zimmerman FJ. Estimation of US Children’s Educational Attainment and Years of Life Lost Associated With Primary School Closures During the Coronavirus Disease 2019 Pandemic. JAMA Netw Open 2020;3:e2028786. DOI:10.1001/jamanetworkopen.2020.28786

11 Viner RM, Mytton OT, Bonell C, et al. Susceptibility to SARS-CoV-2 Infection Among Children and Adolescents Compared With Adults. JAMA Pediatr Published Online First: 25 September 2020. DOI:10.1001/jamapediatrics.2020.4573

12 Aps M, S F. Children are not COVID-19 super spreaders: time to go back to school. Arch Dis Child 2020;105. DOI:10.1136/ARCHDISCHILD-2020-319474

13 Kim J, Choe YJ, Lee J, et al. Role of children in household transmission of COVID-19. Arch Dis Child Published Online First: 7 August 2020. DOI:10.1136/ARCHDISCHILD-2020-319910

14 Stein-Zamir C, Abramson N, Shoob H, et al. A large COVID-19 outbreak in a high school 10 days after schools’ reopening, Israel, May 2020. Eurosurveillance 2020;25:2001352. DOI:10.2807/1560-7917.ES.2020.25.29.2001352

15 Szablewski CM, Chang KT, Brown MM, et al. SARS-CoV-2 Transmission and Infection Among Attendees of an Overnight Camp — Georgia, June 2020. MMWR Morb Mortal Wkly Rep 2020;69:1023–5. DOI:10.15585/mmwr.mm6931e1

16 Larosa E, Djuric O, Cassinadri M, et al. Secondary transmission of COVID-19 in preschool and school settings in northern Italy after their reopening in September 2020: a population-based study. Eurosurveillance 2020;25:2001911. DOI:10.2807/1560-7917.ES.2020.25.49.2001911

17 Otte im Kampe E, Lehfeld A-S, Buda S, et al. Surveillance of COVID-19 school outbreaks, Germany, March to August 2020. Eurosurveillance 2020;25:2001645. DOI:10.2807/1560-7917.ES.2020.25.38.2001645

18 Ismail SA, Saliba V, Bernal JL, et al. SARS-CoV-2 infection and transmission in educational settings: a prospective, cross-sectional analysis of infection clusters and outbreaks in England. Lancet Infect Dis;0. DOI:10.1016/S1473-3099(20)30882-3

19 Buonsenso D, Rose C De, Moroni R, et al. SARS-CoV-2 infections in Italian schools: preliminary findings after one month of school opening during the second wave of the pandemic. medRxiv 2020;:2020.10.10.20210328. DOI:10.1101/2020.10.10.20210328

20 Ulyte A, Radtke T, Abela IA, et al. Seroprevalence and immunity of SARS-CoV-2 infection in children and adolescents in schools in Switzerland: design for a longitudinal, school-based prospective cohort study. Int J Public Health Published Online First: 2020. DOI:10.1007/s00038-020-01495-z

21 West EA, Anker D, Amati R, et al. Corona Immunitas: study protocol of a nationwide program of SARS-CoV-2 seroprevalence and seroepidemiologic studies in Switzerland. Int J Public Health 2020;:1–20. DOI:10.1007/s00038-020-01494-0

22 Institute of Medical Virology IMV SARS-CoV-2 Antikörper Differenzierung (ABCORA).

23 Dan JM, Mateus J, Kato Y, et al. Immunological memory to SARS-CoV-2 assessed for greater than six months after infection. bioRxiv 2020;:2020.11.15.383323. DOI:10.1101/2020.11.15.383323

24 Canton of Zurich. Numbers and Facts on COVID-19 [Kanton Zürich. Zahlen & Fakten zu COVID-19]. https://www.zh.ch/de/gesundheit/coronavirus/zahlen-fakten-covid-19.html?keyword=covid19#/home (accessed 13 Nov 2020).

25 R Core Team. R: A language and environment for statistical computing. R Foundation for Statistical Computing, Vienna, Austria. 2020.https://www.r-project.org/ (accessed 17 Dec 2020).

26 Stan Development Team. RStan: the R interface to Stan. R package version 2.21.2. 2020.

27 Roser M, Ritchie H, Ortiz-Ospina E, et al. Coronavirus Pandemic (COVID-19). 2020.

28 Macartney K, Quinn HE, Pillsbury AJ, et al. Transmission of SARS-CoV-2 in Australian educational settings: a prospective cohort study. Lancet Child Adolesc Heal 2020;0. DOI:10.1016/s2352-4642(20)30251-0

29 Haug N, Geyrhofer L, Londei A, et al. Ranking the effectiveness of worldwide COVID-19 government interventions. Nat Hum Behav 2020;4:1303–12. DOI:10.1038/s41562-020-01009-0

30 Brauner JM, Mindermann S, Sharma M, et al. Inferring the effectiveness of government interventions against COVID-19. Science (80-) 2020;:eabd9338. DOI:10.1126/science.abd9338

31 Auger KA, Shah SS, Richardson T, et al. Association Between Statewide School Closure and COVID-19 Incidence and Mortality in the US. JAMA 2020;324:859. DOI:10.1001/jama.2020.14348

32 Tupper P, Colijn C. COVID-19’s unfortunate events in schools: mitigating classroom clusters in the context of variable transmission. medRxiv 2020;:2020.10.20.20216267. DOI:10.1101/2020.10.20.20216267

33 COVID-19 Schools Infection Survey Round 1, England - Office for National Statistics. https://www.ons.gov.uk/peoplepopulationandcommunity/healthandsocialcare/conditionsanddiseases/bulletins/covid19schoolsinfectionsurveyround1england/november2020 (accessed 19 Dec 2020).

34 Watson J, Whiting PF, Brush JE. Interpreting a covid-19 test result. BMJ. 2020;369. DOI:10.1136/bmj.m1808

